# Auto-detection of motion artifacts on CT pulmonary angiograms with a physician-trained AI algorithm

**DOI:** 10.1101/2022.06.23.22276818

**Authors:** Giridhar Dasegowda, Bernardo C Bizzo, Parisa Kaviani, Lina Karout, Shadi Ebrahimian, Subba R Digumarthy, Nir Neumark, James Hillis, Mannudeep K Kalra, Keith J Dreyer

**Author notes:** Corresponding author: Dr. Bernardo C Bizzo.

## Abstract

**Purpose:** Motion-impaired CT images can result in limited or suboptimal diagnostic interpretation (with missed or miscalled lesions) and patient recall. We trained and tested an artificial intelligence (AI) model for identifying substantial motion artifacts on CT pulmonary angiography (CTPA) that have a negative impact on diagnostic interpretation.

**Methods:** With IRB approval and HIPAA compliance, we queried our multicenter radiology report database (mPower, Nuance) for CTPA reports between July 2015 - March 2022 for the following terms: “motion artifacts,” “respiratory motion,” “technically inadequate,” and “suboptimal” or “limited exam.” All CTPA reports belonged to two quaternary (Site A, n= 335; B, n= 259) and a community (C, n= 199) healthcare sites. A thoracic radiologist reviewed CT images of all positive hits for motion artifacts (present or absent) and their severity (no diagnostic effect or major diagnostic impairment). Coronal multiplanar images belonging to 793 CTPA exams were de-identified and exported offline into an AI model building prototype (Cognex Vision Pro, Cognex Corporation) to train an AI model to perform two-class classification (“motion” or “no motion”) with data from the three sites (70% training dataset, n= 554; 30% validation dataset, n= 239). Separately, data from Site A and Site C were used for training and validating; testing was performed on the Site B CTPA exams. A 5-fold repeated cross-validation was performed to evaluate the model performance with accuracy and receiver operating characteristics analysis (ROC).

**Results:** Among the CTPA images from 793 patients (mean age 63 ± 17 years; 391 males, 402 females), 372 had no motion artifacts, and 421 had substantial motion artifacts. The statistics for the average performance of the AI model after 5-fold repeated cross-validation for the two-class classification included 94% sensitivity, 91% specificity, 93% accuracy, and 0.93 area under the ROC curve (AUC: 95% CI 0.89-0.97).

**Conclusion:** The AI model used in this study can successfully identify CTPA exams with diagnostic interpretation limiting motion artifacts in multicenter training and test datasets.

**Clinical relevance:** The AI model used in the study can help alert the technologists about the presence of substantial motion artifacts on CTPA where a repeat image acquisition can help salvage diagnostic information.

## Introduction

The chest represents one of the most frequently scanned body parts in CT, but also ranks high among the most challenging parts to obtain an optimal diagnostic quality. Recent technical advancements in multidetector-row CT scanners have led to tremendous improvements in diagnostic quality with lower noise, higher contrast, and fewer motion artifacts. Although with faster scanning times and better reconstruction techniques, most chest CT exams are generally optimal for diagnostic interpretation, several artifacts can still have a negative impact on diagnostic interpretations of chest CT [1]. The common artifacts include beam hardening, photon starvation, partial volume, metal, motion, and cone-beam artifacts [2, 3]. Such artifacts can hinder the optimal evaluation of lung abnormalities as well as mediastinal and vascular findings.

Prior studies have reported that motion artifacts are frequent, especially on legacy scanners, and can limit the diagnostic information from both routine chest CT as well as CT pulmonary angiography (CTPA). Motion artifacts can result in misinterpretation as the artifacts can mimic an embolus, or the artifact can cause an apparent abrupt vessel cut-off [4, 5]. In the lung parenchyma, artifacts can reduce the ability to detect and characterize both focal (such as lung nodules) and diffuse parenchymal processes. Such artifacts are especially common in CTPA of critically ill patients and patients with shortness of breath and/or persistent cough [6]. The use of wide-area detector scanners and scan capabilities such as high non-overlapping pitch and faster rotation time reduce scanning duration and have lower artifacts [3, 7]. Such fast acquisition modes are not compatible with dual-energy CT and in patients with large body habitus. Furthermore, such scanners and advanced techniques still represent a minority of clinically deployed scanners even in developed countries such as the United States [8].

An automated method of detecting substantial motion artifacts at the time of scanning can help re-scan patients during the same imaging session with better coaching or faster scanning techniques. Recent advances in machine and deep learning (DL) have led to the creation of several AI algorithms in medical imaging including in the areas of image reconstruction, triaging, quality control and pathology detection [9-14]. Therefore, we trained and tested an artificial intelligence (AI) model to identify substantial motion artifacts on CTPA that have a negative impact on diagnostic interpretation.

## Materials and Methods

### Study Design

Our retrospective study was conducted after receiving approval from Institutional Review Board (IRB). The study was Human Insurance Portability and Accountability Act (HIPAA) compliant. The study methodology has been described in accordance to CLAIM guidelines [15]. The model was trained and tested by physicians without any prior knowledge of, or training in machine learning or coding.

### Data Definitions

The study included adult patients (≥19 years) who underwent CTPA at one of the three hospitals (quaternary hospitals: Massachusetts General Hospital, Brigham and Women’s Hospital; community hospital: Cooley Dickinson Hospital) within an integrated health system. A commercial radiology report search engine, Nuance mPower, was used to identify radiology reports with a mention of motion artifacts in CTPA examinations performed between Jan 2015 to November 2021. The following keywords were used to identify the eligible CTPA: “motion artifacts,” “respiratory motion,” “technically inadequate,” and “suboptimal” or “limited exam.” The search was optimized to identify positive CTPA with the keyword hits (without mention of “absence” or “no” within a few words of the keywords). A negative search with the exact keywords was used to identify the control CTPA exams without motion artifacts.

All CTPA exams were performed on one of the 28 CT scanners in the three participating sites with single or dual-energy CT protocols using the standard of care scan protocols. For each CTPA, a thin slice coronal multiplanar image at the level of descending thoracic aorta was de-identified and exported offline from the PACS workstation.

### Ground truth

In addition to the radiology reports, a thoracic radiologist (MKK with 16 years of subspecialty experience) reviewed all CTPA and opined on the presence or absence of substantial motion artifacts. Each CTPA thus had the opinions of two radiologists, the reporting radiologist, and the study coinvestigator radiologist. Substantial motion artifacts were defined as CTPA exams with the presence of motion artifacts involving both lungs (at least 50% of each lung) and limiting the ability to assess pulmonary embolism or parenchyma. Minor motion artifacts involving a single lobe or smaller portions of lungs without effect on the diagnostic evaluation of pulmonary embolism or parenchyma were labeled as negative for substantial motion artifacts. Although extremely common, minor artifacts are not as important since they should not trigger repeat imaging. CTPA with evidence of “white lungs” (diffuse parenchymal opacities), a substantial bilateral lung volume loss, or pneumonectomies were excluded from the study (n= 50 CTPA exams). These cases were excluded since it is difficult to assess motion artifacts’ impact on the pulmonary evaluation in such cases. CTPA exams without the complete inclusion of lung apex and bases were excluded from the study.

### Model

The AI model was trained on a deep learning model-building platform, Vision Pro Deep Learning (VPDL, COGNEX Corporation, Natick, MA). The software enables users to train DL models based on a labeled image dataset using a vision-optimized deep neural network. The users require no formal programming or coding knowledge or experience. The VPDL platform has two different options for training classification models: High detail Mode (HDM) and Focused Mode (FM). The HDM enables model training for challenging or complex applications (such as pixel-level information in image domain) and provides higher accuracy. A heat map is also generated in HDM that indicates the image region that was most influential in the classification decision. FM enables fast training of models for simple applications (such as the distinction between different image types or body parts). We used the HDM classification model for identifying CTPA examinations with substantial motion artifacts.

### Training

A physician co-investigator (GD with one-year post-doctoral research fellowship experience in thoracic imaging) trained the AI model on the VPDL platform without prior programming or data science knowledge. The images were de-identified, exported from the PACS workstation (Visage), and then uploaded onto the software platform which was installed on a virtual machine within the hospital intranet to maintain data security and privacy. Within the platform, the study coinvestigator labeled each uploaded image as “motion” (with substantial motion artifacts) or “no motion” (without substantial motion artifacts) based on the assessment from the thoracic radiologist. In the first training, all CTPA examinations from three hospitals were included. The software randomized the training and validation data set with a 70%-30% distribution, respectively. We performed five-fold repeated cross-validation to evaluate the robustness of the model. The output was recorded and separately analyzed.

To establish the inter-institutional generalizability of the model, we trained a model using images from two hospitals (A and C, after excluding the data from site B), and then tested the algorithm on CTPA data from the third hospital (site B).

### Statistical Analysis

Information on the distribution of true positive, true negative, false positive, and false negative CTPA was recorded in Microsoft Excel worksheets (Microsoft Inc). Data were analyzed with SPSS statistical software, version 26 (IBM Inc.). The performance of the AI model was evaluated using sensitivity, specificity, accuracy, and area under the curve (AUC) for the receiver operating characteristic (ROC) analysis. For the five-fold repeated cross-validation, the average of the five models was considered. We also estimated the F-score for AI model performance, as a measure of the harmonic average of precision and recall/sensitivity.

## Results

Our study included 793 CTPA examinations from 793 adult patients (mean age= 63 ± 17; 391 men, 402 women). The distribution of CTPA across each site is as follows: Site A, n= 335; Site B, n= 259; Site C, n= 199. A total of 455/793 (57%) CTPA examinations were performed during the emergency visit, while 277/793 (35%) and 111/793 (14%) examinations were among inpatients and outpatients, respectively. Most CTPA with substantial motion artifacts were either from the emergency department (n = 213/455) or inpatients (n = 146/227) with a minority of patients coming with outpatient referrals (n = 33/111). There was no discrepancy between the reporting radiologists and the research radiologist for the presence of substantial motion artifacts.

### Model validation at three sites

Among CTPA exam datasets from all three sites in model training, 471 CTPA (n= 421/793, 53.1%) had substantial motion artifacts and 372 CTPA (n= 372/793, 46.9%) were without substantial artifacts. For the average performance after 5-fold repeated cross-validation for the two-class classification (as either with or without substantial motion artifacts), on the 30% validation data set, the AI model had a sensitivity of 94%, specificity of 91%, 93% accurate with AUC of 0.93 (95% confidence interval [95% CI] 0.89 - 0.97). The best performing model had F-scores of 96% and 95% for identifying CTPA with and without substantial motion artifacts, respectively. The two-class classification of the AI model is summarized in Fig 1.

**Fig 1.**
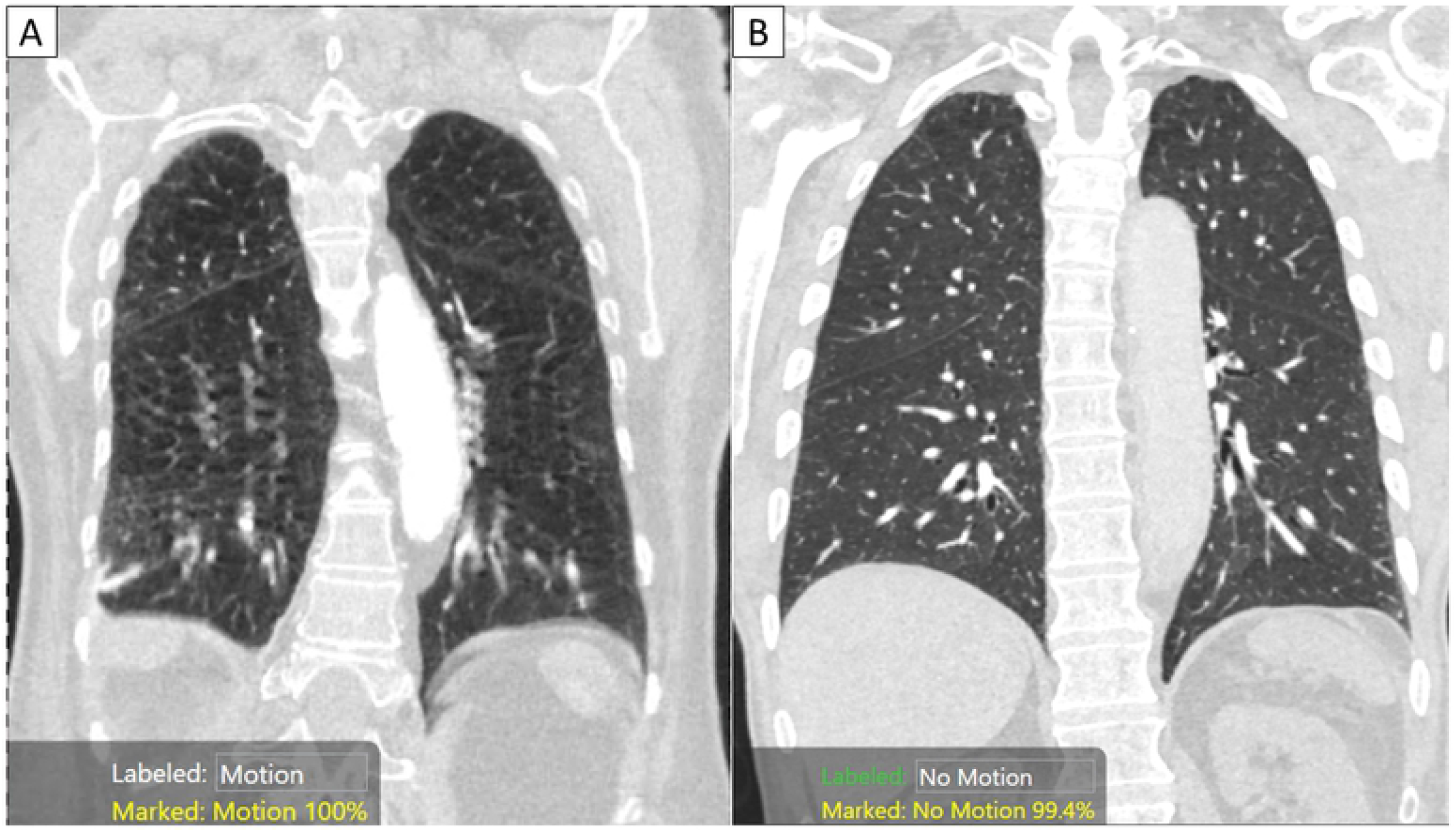
Coronal MPR images of CTPA examinations in two patients with (A) and without substantial motion artifacts. The AI algorithm correctly classified the images as with motion (A: with 100% confidence score) and “no motion” (B: with 99.4% confidence score).

### Model testing

External testing with Sites A and C training datasets and Site B as the test site, the model performance statistics were 85% sensitivity, 90% specificity, 86% accuracy, and an AUC of 0.87 (95% CI 0.82 -0.92). One hundred forty-seven CTPA exams were correctly classified into those with substantial motion artifacts (true positive), and nine CTPA were mislabeled as positive for motion artifacts (false positive). Seventy-seven CTPA exams were correctly classified as without motion artifacts (true negative), and twenty-six CTPA exams were misclassified as without motion (false negative). The confusion matrix of model performance is represented in Fig 2 and the model performance for validation and test data is summarized in Table 1.

**Fig 2:**
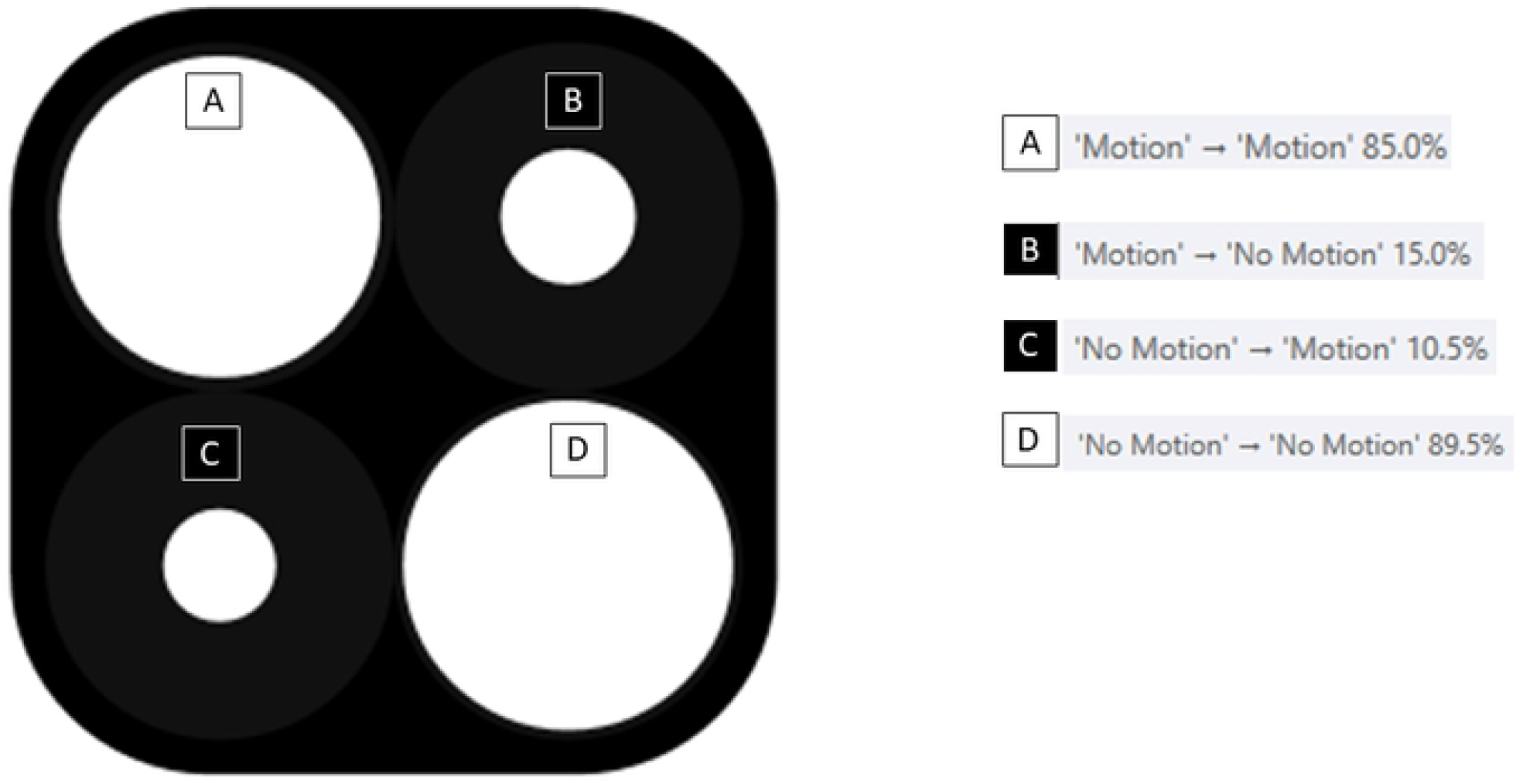
Confusion matrix generated by the AI model for the testing (A true positive rate [sensitivity] – AI correctly labeled images with motion; B false-negative rate – AI incorrectly labeled “motion” images as without motion; C false positive rate – AI incorrectly labeled “no motion” images as with substantial motion artifacts; D true negative rate [specificity]– AI correctly identified images without motion artifacts).

**Table 1.** Summary of our AI model’s performance for motion artifacts detection in CTPA exams from the three participating sites (A, B, C). (key: + present; -absent; CI confidence interval).

|  |  | Training | Test | Sensitivity | Specificity | Accuracy | AUC | 95% CI (AUC) |
| --- | --- | --- | --- | --- | --- | --- | --- | --- |
| Model validation<br>(training + validation<br>data from all sites) | Motion + | 295 | 126 | 94% | 91% | 93% | 0.93 | 0.89-0.97 |
|  | Motion - | 259 | 113 |  |  |  |  |  |
| Model testing<br>(training from A, C<br>and testing on B) | Motion + | 196 | 173 | 85% | 90% | 86% | 0.87 | 0.82-0.92 |
|  | Motion - | 231 | 86 |  |  |  |  |  |

## Discussion

We report high accuracy, sensitivity, and specificity of physician-trained and tested AI model for identifying substantial motion artifacts on CTPA examinations. Prior studies have reported on the ability of AI algorithms to identify anatomic regions with motion artifacts [16]. Our study uses a single coronal multiplanar reformatted image per CT exam and therefore could be more time-efficient, while ignoring region-specific, sparse motion artifacts which do not require repeat acquisition [17-19]. To our best knowledge, there are no peer-reviewed reports on the use of AI models for identifying motion artifacts in CTPA or chest CT examinations.

Beri *et al*. reported that their AI algorithm trained to identify the motion affected regions by segmenting the entire image series had an AUC of 0.81 [18]. We achieved a similar AUC of 0.88 with a single coronal MPR image per CTPA examination. Other studies [17, 19] on motion artifact detection in CT images have focused in coronary CT angiography (CCTA) rather than CTPA examinations. Ma *et al*. reported 91% sensitivity and 71% specificity with 87% accuracy for detecting motion artifacts in CCTA examinations [17]. Likewise, Elss *et al*. trained an AI algorithm for identifying motion artifacts on CCTA and reported an accuracy of 94% [19]. Xu *et al*. reported a fully automatic AI for grading image quality (motion artifacts) of CCTA using semi-automatic labeling and tracking of the coronary arteries [20] Based on the identification and estimation of motion artifact, other investigators have reported on motion artifact correction and compensation solutions for head and cardiac CT examinations [21, 22].

Although not yet cleared by the US Food and Drug Administration (FDA), our proof-of-concept study and the AI model used in the study may have potential clinical implications. Firstly, given the high frequency of motion artifacts in chest CT and CTPA examinations, our study highlights the role of AI models in identifying motion artifacts. If integrated with CT scanners, such AI models can efficiently detect and alert the CT technologist to the presence of motion artifacts that are likely to have a substantial effect on the diagnostic interpretation. Secondly, such artifacts can be present on chest CT exams acquired on both the older, less advanced scanners and newer, faster scanners. On the latter, modification of scanning parameters can enable faster scanning of the entire chest in under 1 second [23]. On the older scanner, the AI-generated surveillance for motion-impaired CTPA or chest CT examinations can prompt technologist to give better breath-hold instructions [24] and modify scan parameters or when possible scan at-risk patients on faster scanners. Thirdly, most diagnostic CT scanners in our institution (MGH) have two scan protocols – one for patients who can hold their breath and the other for those who are unable to hold their breath or have substantial motion artifacts on their initial CT acquisition. Despite instructions to our CT technologists to always review the CT images for motion artifacts before taking the patient off the CT table, most technologists cannot or do not comply with the recommendation especially during pandemic times. As a result, interpreting radiologists either report CT with a disclaimer on motion-limited diagnostic value or request patient recall and rescanning. By automating the detection of motion artifacts, AI models such as the one reported in our study could potentially help address compliance and reacquisition, when appropriate. Fourthly, several modern scanners automatically generate multiplanar reformatted images as soon as the data acquisition is complete, so the use of coronal MPR image for our model is not a rate-limiting step. Although our model would still require the identification of the single image at the descending aorta level. Finally, in hospitals with high CT volumes, the task of quality assessment for image quality is time-consuming and labor-intensive. In such sites, the AI model used in this study can be used to analyze image quality in a retrospective manner. Derived statistics can then be used to develop faster scan protocols and track their impact on diagnostic evaluability. Our study has limitations. We did not perform a power analysis to determine the number of training and testing cases needed to prove our hypothesis. Although the high level of performance associated with the AI model suggests that our sample size was adequate, it is conceivable that more training data could generate better results. Likewise, for external testing, we had only a single site with completely independent clinical operations, and we would require additional external sites to support the claim of generalizability of the AI model. The AI model’s performance can however vary with the change in scan protocols (low dose chest CT versus CTPA protocols) and scanners (for those scanners without input training data). Exclusion of CTPA with “white lungs” (diffuse parenchymal opacities), a substantial bilateral lung volume loss, or pneumonectomies also limits the application of our AI model in such patients.

Another limitation of our study pertains to the use of a single coronal MPR image per CT for assessing substantial motion artifacts as opposed to the entire image series for prior studies [18]. It is therefore possible that the performance of our AI model can differ on the entire image series (transverse or coronal) as compared to its current performance on a single image. Despite a high model performance, it is possible that motion artifacts in other anatomic location can affect evaluation of key findings. Given the full longitudinal coverage of anatomy in the coronal plane, we believe that such “missed motion artifacts in key location” is less likely.

The AI building platform at the time of manuscript preparation cannot easily group entire image series and is limited to 2D image input, which impacted our decision to train on the coronal MPR image instead of an axial series. Finally, our model training and testing were limited to CTPA and might not apply to other chest CT protocols or body regions.

In conclusion, the physician-trained and tested AI model can help identify substantial motion artifacts on CT pulmonary angiography. Automatic recognition of such artifacts can help CT technologists apply faster scan protocols and reacquire images to mitigate the impact of substantial motion impairment on diagnostic evaluability.

## Data Availability

All relevant data are within the manuscript and its Supporting Information files.

